# Person-centeredness of the San Francisco Pregnancy Village model of cross-sector care delivery: A mixed-methods study

**DOI:** 10.1101/2025.06.05.25329089

**Authors:** Prisca C. Diala, Osamuedeme J. Odiase, April J. Bell, Alison M. El Ayadi, KaSelah Crockett, Malini A. Nijagal, Patience A. Afulani

## Abstract

**Background:** Racial disparities in perinatal outcomes are worsened by systemic racism in the United States. San Francisco’s Pregnancy Village is a novel cross-sector care delivery model providing a one-stop-shop for city government, clinical, and community-based wraparound services, delivered in an uplifting, celebratory, and dignified environment to center Black and other minoritized pregnant people and their families.

**Objectives:** To evaluate the person-centeredness of the Pregnancy Village model’s first event series: the SF Family & Pregnancy Pop-Up Village.

**Design:** We employed a convergent mixed-methods study design. Data were collected between July 2021 and June 2022.

**Methods:** We conducted quantitative surveys with 116 Pregnancy Village participants (57 pregnant/postpartum individuals and 59 family members) and semi-structured in-depth interviews with purposively sampled 13 pregnant/postpartum people and five family members. Person-centered care (PCC) was assessed using a 10-item scale. Standardized scores ranged from 0 to 100, where higher scores reflect higher PCC. We performed univariate, bivariate, and multivariate analyses, and thematic analyses of the qualitative data.

**Results:** The mean PCC score was 91.2 (SD = 12.1). Participants who experienced discrimination during prenatal encounters had significantly lower PCC scores than those who did not. In general, participants perceived the Pregnancy Village events as person-centered. A welcoming environment, patient visibility, and warm handoffs reflected responsive and supportive care; provider friendliness and intentionality reflected dignity and respect; and provider knowledge, inclusivity, and genuineness represented communication and autonomy. Identified gaps were related to follow-up and communication.

**Conclusion:** Our findings indicate that the Pregnancy Village model is person-centered and enables pregnant people and their families to receive care that prioritizes their needs, preferences, and values and has the potential to reduce health inequities in care experience and outcomes. Close follow-up after such events and providing language concordant support is crucial to achieving model goals.

## INTRODUCTION

With 22.3 maternal deaths per 100,000 live births, the United States (U.S.) has the highest rate of maternal mortality of any high-income country (1). However, the burden of mortality falls disproportionately on Black women and birthing people. In 2022, the rate of maternal mortality among Black non-Hispanic individuals was 49.5 deaths per 100,000 live births, more than double the ratio observed among Latine, Asian, and White individuals^1^ (1). Black non-Hispanic women also face a 70% greater risk of severe maternal morbidity (3) and 50 percent higher rates of preterm birth than White or Hispanic individuals (4), contributing to significant perinatal health disparities.

As perinatal disparities and maternal mortality and morbidity increase, addressing inequities in care and perinatal health outcomes, including preterm birth and infant death, is urgent. Recent literature has identified an inverse relationship between structural and interpersonal racism and quality of healthcare delivery, providing robust evidence that experiences of racism result in adverse outcomes (5–7). Many pregnant individuals of color report feelings of dismissal, judgment, and lack of autonomy in their interactions with healthcare providers, which in turn disrupts the patient-provider alliance and undermines trust, which is crucial for positive care experiences and the provision of high-quality, effective care (8–10). In this context, models of care that aim to reduce structural and interpersonal racism while fostering trust between pregnant individuals and care providers are crucial. These models should prioritize personalized, accessible, and culturally competent care that addresses the unique needs of Black individuals, thus playing a vital role in tackling health inequities (11).

Recognizing that current U.S. systems for providing healthcare and support are a root cause of inequities (12), our team embarked on a year-long Human-Centered Design (HCD) process in 2017 to address perinatal inequities among Black birthing individuals in San Francisco. Our findings confirmed extant evidence that U.S. health and social systems present significant practical barriers to access, are not trusted by communities of color, and fail to honor the dignity and autonomy of Black pregnant individuals (13). This process led to the “Pregnancy Village” model of improving care access and experience by providing a “one-stop-shop” of cross-sector services in an environment designed to be dignified and celebratory and prioritizing trust-building and holistic health and wellness by addressing the comprehensive needs of individuals, families, and communities (14). Following a three-year planning phase, the first iteration was launched in collaboration with the “Pop-Up Village” model founders, Designing Justice + Designing Spaces: the SF Family & Pregnancy Pop-Up Village event series (subsequently referred to as the “Pregnancy Village” (PV) for brevity).

We sought to comprehensively evaluate the implementation of the Pregnancy Village model, including feasibility, fidelity, participant perceptions of acceptability, and preliminary effects on accessibility, person-centeredness, comfort, and trustworthiness. This paper focuses on the person-centeredness of the model. Person-centered care (PCC), which prioritizes meeting individuals’ needs, preferences, and values in a manner that is responsive and respectful (Figure 1) (15,16), is an important tenet of the model and an avenue through which care systems and providers may successfully rebuild the patient-provider alliance. A robust understanding of person-centeredness at events can inform our program’s goal to optimize the model to ensure acceptability and responsiveness to community needs through continual community engagement and work with providers outside events. Therefore, the present analysis seeks to understand participant perspectives of person-centered care of the PV events and factors associated with person-centered care, concentrating on the most formative phase: the first nine monthly events (July 2021-June 2022).

**Figure 1.**
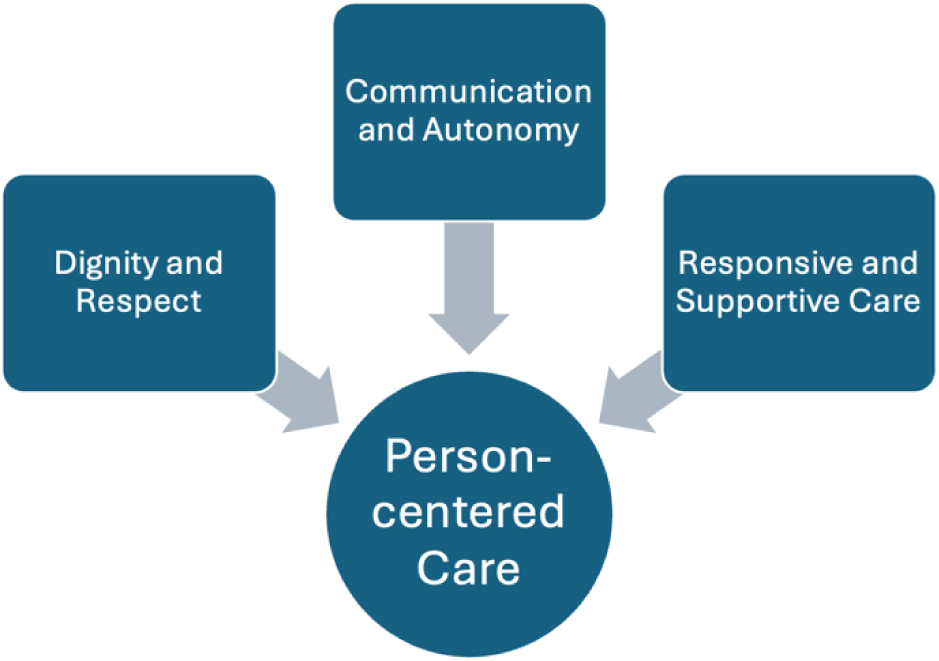
Domains of person-centered care.

## METHODS

### Setting

Briefly, PV events took place in San Francisco’s Bayview district, home to nearly 40 percent of the city’s Black birthing residents and nearly 20 percent of its publicly insured birthing residents (17). Within the Bayview, 61% of all birthing people are Medicaid-insured, and 93% identify as members of a racial or ethnic minority group (18). Compared to other neighborhoods, Bayview residents have significantly higher rates of preterm birth and lower rates of timely prenatal care, highlighting perinatal care inequities (18).

### Intervention

The Pregnancy Village model of cross-sector collaboration seeks to reduce perinatal inequities by providing a one-stop-shop for comprehensive care and support in a celebratory and uplifting environment designed for Black pregnant people and their families. Events occur monthly and feature various participating partners, including city government, healthcare, and community-based organizations. Offerings range from traditional health services (e.g., Medicaid enrollment and consultations with a healthcare provider) to comprehensive wellness services (e.g., dance and food preparation classes, massage, acupuncture, and sharing circles). The model is grounded in anti-racism and person-centered principles, emphasizes sustainable community-institutional partnerships, and incorporates a real-time community feedback system for responsive model iteration. Further details about the first implementation of the model have been described elsewhere (14).

### Study design

We employed a convergent mixed methods design where quantitative and qualitative data from surveys and in-depth interviews were collected concurrently, analyzed separately, and then integrated to interpret the results (19). Integration also occurred by sampling a subset of the survey participants for the in-depth interviews and developing the in-depth interview guide to expand on questions from the survey. The study findings are reported in accordance with The Good Reporting of a Mixed Methods Study (GRAMMS) (20) and the Consolidated Criteria for Reporting Qualitative Research (COREQ) guidelines (21).

### Sample

While events were designed to tackle inequities faced by Black pregnant and postpartum individuals and their families, the organizers believed that the offerings could also benefit other community members facing barriers to health, so events were made inclusive. We therefore recruited Black and other minoritized pregnant and postpartum people and their family members who participated in at least one of the nine monthly events from July 2021 to June 2022 through convenience sampling. The eligibility criteria were as follows: 1) individuals who were pregnant or postpartum (having given birth within the past year) and were at least 15 years old; 2) family members who were at least 18 years old; 3) individuals who spoke English or Spanish; and 4) participated in at least one of the nine monthly events.

### Quantitative data collection

The target quantitative sample was 120 participants based on feasibility— anticipating that we could recruit 10-15 individuals at each event. Potential participants were asked during event registration whether they would be interested in learning about a study to evaluate their experience. Those who expressed interest were approached and screened by the evaluation team. Eligible individuals provided verbal informed consent after learning about the study. Participants were requested to complete a survey regarding their experience onsite using a tablet or later on their own devices using a QR code. Surveys were self-administered (except in a few cases per participant request) and completed in English or Spanish. Participants were permitted to complete the survey more than once if they attended multiple PV events, and fifteen individuals did so.^2^ The survey assessed various aspects of the PV experience, notably the person-centeredness of PV. All participants were compensated with a $20 gift card.

### Qualitative data collection

A subset of the survey participants (n=18) was purposively sampled for semi-structured in-depth interviews, maintaining a ratio of 70% of pregnant and postpartum individuals (n=13) to 30% of family members (n = 5). Survey participants were asked if they would be interested in participating in a follow-up interview, and three to four individuals from each PV event were purposively selected and invited onsite or contacted by phone to participate in in-depth interviews. These interviews were scheduled at a convenient time for the participants. A semi-structured interview guide was developed to cover key PCC domains. The interviews lasted between 30 to 60 minutes and were held via Zoom in either English or Spanish. They occurred within four weeks of the participants attending a PV event and were conducted by researchers with qualitative training (OJO and KV). Participants received an additional $20 for their participation in the interviews. With participants’ permission, interviews were recorded and transcribed by a third-party transcriptionist. Research assistants (KV and JV) verified the accuracy and clarity of the transcripts. Field notes were taken during interviews, and a standardized template was developed afterward to facilitate rapid analysis for model iteration.

### Quantitative Measures

The primary outcome of the quantitative analysis, the PCC score, was measured using a set of items adapted from the person-centered prenatal care (PCPC) scale. The PCPC scale was developed to capture the prenatal experiences of racial and ethnic minority groups and validated among Black and Latinx women (16,22). The adapted scale comprised ten close-ended questions spanning the PCC domains of dignified and respectful care, communication and autonomy, and responsive and supportive care (Appendix 1). From the four-point frequency response options [i.e., 0-(“No, not at all”), 1-(“A little”), 2-(“Somewhat”), 3-(“Yes, definitely”)], we generated a summative score from the responses to the individual items. Negatively worded questions were reverse-coded, and not applicable response options were coded to the upper middle category before generating summative scores (16,22). Missing data (2.7%) were imputed as the mean of other items in the measure. The scores were standardized on a scale of 0 to 100 for ease of interpretation. The Raykov’s rho for the full sample was 0.71.

### Covariates

Participants were asked to report on various sociodemographic factors, such as race and ethnicity, age, gender, level of education, proficiency in English, preferred language, neighborhood of residence, housing status, social support, relationship status, medical insurance status, employment status, receipt of public income assistance, and food insecurity (23) (see Table 1). Additionally, participants reported on various obstetric factors, including their pregnancy status (either pregnant or postpartum, or family member of a pregnant or postpartum individual), parity, history of pregnancy loss (e.g., induced abortion, miscarriage, or stillbirth), history of preterm birth, and prenatal care attendance during their current or recent pregnancy (if they were pregnant or postpartum). The survey also included questions about previous experiences of discrimination, which were adapted from the Everyday Discrimination Scale and Discrimination in Medical Settings Scale (24,25).

**Table 1.** Univariate distribution of sociodemographic characteristics, obstetric history, and care discrimination experiences among currently/recently pregnant and family participants in the San Francisco Family and Pregnancy Pop-Up Village, San Francisco, CA.

|  | **Total (N=116)** | **Currently pregnant/recently pregnant (n=57)** | **Neither pregnant nor recently pregnant (n=59)** |
| --- | --- | --- | --- |
| **Participant Characteristics** | **% (N)** | **% (n)** | **% (n)** |
| *Age* |  |  |  |
| 15-24 | 11.2% (13) | 17.5% (10) | 5.1% (3) |
| 25-34 | 26.7% (31) | 38.6% (22) | 15.3% (9) |
| 35-44 | 21.6% (25) | 33.3% (19) | 10.2% (6) |
| 45 and older | 25.9% (30) | 1.8% (1) | 49.2% (29) |
| Unknown | 14.7% (17) | 8.8% (5) | 20.3% (12) |
| *Gender* |  |  |  |
| Female | 86.2% (100) | 91.2% (52) | 81.4% (48) |
| Male | 5.2% (6) | 0.0% (0) | 10.2% (6) |
| Other/unknown/prefer not to answer | 8.6% (10) | 8.8% (5) | 8.5% (5) |
| *Race/ethnicity* |  |  |  |
| Non-Hispanic Black | 40.5% (47) | 26.3% (15) | 54.2% (32) |
| Hispanic/Latine | 36.2% (42) | 47.4% (27) | 25.4% (15) |
| Multiracial | 15.5% (18) | 19.3% (11) | 11.9% (7) |
| Other/unknown/prefer not to answer | 7.8% (9) | 7.0% (4) | 8.5% (5) |
| *Language* |  |  |  |
| English | 59.5% (69) | 52.6% (30) | 66.1% (39) |
| Spanish | 30.2% (35) | 38.6% (22) | 22.0% (13) |
| Unknown/other/prefer not to answer | 10.3% (12) | 8.8% (5) | 11.9% (7) |
| *English proficiency* |  |  |  |
| Very well or well | 70.7% (82) | 70.2% (40) | 71.2% (42) |
| With difficulty | 17.2% (20) | 19.3% (11) | 15.3% (9) |
| Unknown/prefer not to answer | 12.1% (14) | 10.5% (6) | 13.6% (8) |
| *Pregnancy status* |  |  |  |
| Family member | 50.9% (59) | 0.0% (0) | 100.0% (59) |
| Pregnant or postpartum | 49.1% (57) | 100.0% (57) | 0.0% (0) |
| *Parity* |  |  |  |
| No births | 22.4% (26) | 26.3% (15) | 18.6% (11) |
| 1-2 births | 43.1% (50) | 47.4% (27) | 39.0% (23) |
| 3 or more births | 24.1% (28) | 24.6% (14) | 23.7% (14) |
| Unknown/not applicable/prefer not to answer | 10.3% (12) | 1.8% (1) | 18.6% (11) |
| *Prenatal care* |  |  |  |
| No prenatal care during current/recent pregnancy | 3.4% (4) | 7.0% (4) | 0.0% (0) |
| Received prenatal care during current/recent pregnancy | 41.4% (48) | 84.2% (48) | 0.0% (0) |
| Unknown/not applicable/ prefer not to answer | 55.2% (64) | 8.8% (5) | 100.0% (59) |
| *History of preterm birth* |  |  |  |
| No preterm births | 69.0% (80) | 80.7% (46) | 57.6% (34) |
| At least 1 preterm birth | 19.8% (23) | 15.8% (9) | 23.7% (14) |
| Unknown/not applicable/prefer not to answer | 11.2% (13) | 3.5% (2) | 18.6% (11) |
| *History of pregnancy loss* |  |  |  |
| No prior pregnancy loss | 59.5% (69) | 57.9% (33) | 61.0% (36) |
| Pregnancy loss | 25.0% (29) | 35.1% (20) | 15.3% (9) |
| Unknown/not applicable/prefer not to answer | 15.5% (18) | 7.0% (4) | 23.7% (14) |
| *Education* |  |  |  |
| Less than a high school degree | 18.1% (21) | 15.8% (9) | 20.3% (12) |
| High school graduate, GED or equivalent | 25.9% (30) | 29.8% (17) | 22.0% (13) |
| Some college, junior college or vocational school | 27.6% (32) | 21.1% (12) | 33.9% (20) |
| College graduate or higher | 17.2% (20) | 21.1% (12) | 13.6% (8) |
| Unknown/prefer not to answer | 11.2% (13) | 12.3% (7) | 10.2% (6) |
| *Employment* |  |  |  |
| No | 63.8% (74) | 56.1% (32) | 71.2% (42) |
| Yes, Full-time | 13.8% (16) | 19.3% (11) | 8.5% (5) |
| Yes, Part-time | 10.3% (12) | 15.8% (9) | 5.1% (3) |
| Unknown/prefer not to answer | 12.1% (14) | 8.8% (5) | 15.3% (9) |
| *Income assistance* |  |  |  |
| No | 43.1% (50) | 33.3% (19) | 52.5% (31) |
| Yes | 47.4% (55) | 54.4% (31) | 40.7% (24) |
| Unknown/prefer not to answer | 9.5% (11) | 12.3% (7) | 6.8% (4) |
| *Residence* |  |  |  |
| Bayview-Hunter's Point | 33.6% (39) | 38.6% (22) | 28.8% (17) |
| Other San Francisco | 44.8% (52) | 43.9% (25) | 45.8% (27) |
| East Bay | 9.5% (11) | 12.3% (7) | 6.8% (4) |
| Other/unknown/prefer not to answer | 12.1% (14) | 5.3% (3) | 18.6% (11) |
| *Housing* |  |  |  |
| Homeless Shelter | 10.3% (12) | 12.3% (7) | 8.5% (5) |
| Living with someone for free | 4.3% (5) | 5.3% (3) | 3.4% (2) |
| No living place | 0.9% (1) | 0.0% (0) | 1.7% (1) |
| Owns home or apartment | 6.9% (8) | 5.3% (3) | 8.5% (5) |
| Public Housing | 14.7% (17) | 12.3% (7) | 16.9% (10) |
| Renting home or apartment | 50.0% (58) | 52.6% (30) | 47.5% (28) |
| Transitional or supportive housing | 2.6% (3) | 3.5% (2) | 1.7% (1) |
| Other/unknown/prefer not to answer | 10.3% (12) | 8.8% (5) | 11.9% (7) |
| *Social Support* |  |  |  |
| Not at all | 12.9% (15) | 7.0% (4) | 18.6% (11) |
| A little | 9.5% (11) | 14.0% (8) | 5.1% (3) |
| Somewhat | 19.0% (22) | 22.8% (13) | 15.3% (9) |
| Yes, definitely | 54.3% (63) | 54.4% (31) | 54.2% (32) |
| Unknown/prefer not to answer | 4.3% (5) | 1.8% (1) | 6.8% (4) |
| *Medical insurance status* |  |  |  |
| Public insurance | 66.4% (77) | 66.7% (38) | 66.1% (39) |
| Private insurance | 15.5% (18) | 19.3% (11) | 11.9% (7) |
| No insurance | 4.3% (5) | 1.8% (1) | 6.8% (4) |
| Unknown/prefer not to answer | 13.8% (16) | 12.3% (7) | 15.3% (9) |
| *Food insecurity (Worried food would run out)* |  |  |  |
| Often true | 11.2% (13) | 8.8% (5) | 13.6% (8) |
| Sometimes true | 24.1% (28) | 33.3% (19) | 15.3% (9) |
| Never true | 48.3% (56) | 36.8% (21) | 59.3% (35) |
| Unknown/prefer not to answer | 16.4% (19) | 21.1% (12) | 11.9% (7) |
| *Food insecurity (Insufficient funds)* |  |  |  |
| Often true | 11.2% (13) | 12.3% (7) | 10.2% (6) |
| Sometimes true | 27.6% (32) | 36.8% (21) | 18.6% (11) |
| Never true | 43.1% (50) | 33.3% (19) | 52.5% (31) |
| Unknown/prefer not to answer | 18.1% (21) | 17.5% (10) | 18.6% (11) |
| *Relationship status* |  |  |  |
| Partnered, living together | 31.9% (37) | 42.1% (24) | 22.0% (13) |
| Partnered, not living together | 13.8% (16) | 17.5% (10) | 10.2% (6) |
| Single | 42.2% (49) | 24.6% (14) | 59.3% (35) |
| Other/unknown/prefer not to answer | 12.1% (14) | 15.8% (9) | 8.5% (5) |
| *Everyday discrimination experience* |  |  |  |
| Never | 17.2% (20) | 19.3% (11) | 15.3% (9) |
| Rarely | 25.9% (30) | 28.1% (16) | 23.7% (14) |
| Sometimes | 36.2% (42) | 35.1% (20) | 37.3% (22) |
| Often | 16.4% (19) | 15.8% (9) | 16.9% (10) |
| Unknown/prefer not to answer | 4.3% (5) | 1.8% (1) | 6.8% (4) |
| *Prenatal care discrimination experience* |  |  |  |
| Never | 42.2% (49) | 31.6% (18) | 52.5% (31) |
| Rarely | 19.0% (22) | 26.3% (15) | 11.9% (7) |
| Sometimes | 22.4% (26) | 35.1% (20) | 10.2% (6) |
| Often | 6.9% (8) | 5.3% (3) | 8.5% (5) |
| Unknown/prefer not to answer | 9.5% (11) | 1.8% (1) | 16.9% (10) |

### Quantitative Data Analyses

Of the 120 participants enrolled, four were deemed ineligible after reviewing their demographic information, resulting in a final analytical sample of 116 participants. Descriptive statistics were employed to assess the sample’s sociodemographic characteristics and the distribution of the PCC scores. Bivariate analyses of predictor and outcome measures were then performed using crosstabulations and ordinary least squares (OLS) regression to examine statistically significant associations. Multivariate models included predictors that were statistically significant (p < 0.05). Multivariate models were refined by removing closely related variables discovered during collinearity tests and model fit assessments. To account for clustering caused by some participants answering more than once, we estimated linear mixed effects models. We also performed sensitivity analyses to evaluate when “Not applicable” responses in the PCC scale were coded as missing, and missing responses were excluded from the analysis together with duplicate responses (subsequent surveys from the same respondents). All analyses were conducted using STATA (version 14), and statistical differences were deemed significant at p<0.05.

### Qualitative data analysis

We utilized a thematic analysis approach to identify themes related to PCC (26). The qualitative lead (OJO) created an initial deductive codebook based on the interview guide. Six analysts (OJO, PD, KV, EK, HS, and KM) coded the transcripts independently using Dedoose software. To ensure inter-rater reliability, two transcripts were collaboratively coded. We conducted a detailed comparison of our coding, refined the codebook, and added inductive codes. The remaining transcripts were coded by balanced pairs of analysts. The qualitative lead reviewed all coding to ensure consistency. We then queried and analyzed codes specific to PCC to identify emerging themes. The analytic summaries of these codes were organized into a thematic table and categorized by PCC sub-domains: 1) dignity and respect, 2) communication and autonomy, and 3) responsive and supportive care.

## RESULTS

### Quantitative findings

#### Participant Characteristics

Table 1 summarizes the sociodemographic and obstetric characteristics of participants. About half (49%) of participants were presently or recently pregnant, with 29% currently pregnant. About 27% were between the ages of 25 and 34, while 22% were between the ages of 35 and 44. Around 41% of participants identified as Black or African American, and 36% as Latine. Eighteen percent had not completed high school, while 45% pursued education beyond high school. About two-thirds (64%) of participants reported being unemployed, with 47% receiving income assistance. Two-thirds of participants had public medical insurance. Forty-two percent of participants were single, while 32% reported living with a romantic partner. Twenty percent of participants reported a history of preterm birth, and one-fourth of participants had a past pregnancy loss.

#### Distribution of PCC

The sample’s standardized mean PCC score was 91.2 (*SD* = 12.1) overall, 90.1 (*SD* = 13.2) for pregnant and postpartum participants, and 91.5 (*SD* = 11.0) among family members, t(111) = 0.277, *p* = 0.790 (see Table 2). The mean PCC score for Black participants only (N = 47) was 91.8 (SD = 12.2), compared to a mean PCC score of 90.7 (SD = 12.2) for participants from all other racial and ethnic groups, t(111) = 0.490, *p* = 0.625.

**Table 2.** Distribution of standardized person-centered care score among currently/recently pregnant and family participants in the San Francisco Family and Pregnancy Pop-Up Village, San Francisco, CA.

|  | *N* | *Score* | *SD* | **Minimum**<br>**Score** | **Maximum**<br>**Score** |
| --- | --- | --- | --- | --- | --- |
| Combined Sample | 113 | 91.2 | 12.1 | 43.3 | 100.0 |
| Pregnant/postpartum | 57 | 90.1 | 13.2 | 43.3 | 100.0 |
| Family member | 56 | 91.5 | 11.0 | 53.3 | 100.0 |
| Black participants | 47 | 91.8 | 12.2 | 43.3 | 100.0 |
| Participants from all other racial/ethnic groups | 66 | 90.7 | 12.2 | 46.7 | 100.0 |

Abbreviations: SD: standard deviation

#### Factors associated with PCC score

In bivariate analyses, participants who preferred not to disclose their residence, did not disclose their medical insurance status, and experienced discrimination during their usual prenatal care encounters on some occasions had lower perceptions of PCC than participants who lived in the Bayview, had public insurance, and never experienced discrimination during prenatal encounters, respectively (Table 3; Appendix 2). Additionally, participants who reported owning a home or apartment had higher perceptions of PCC than those who reported renting.

**Table 3.**
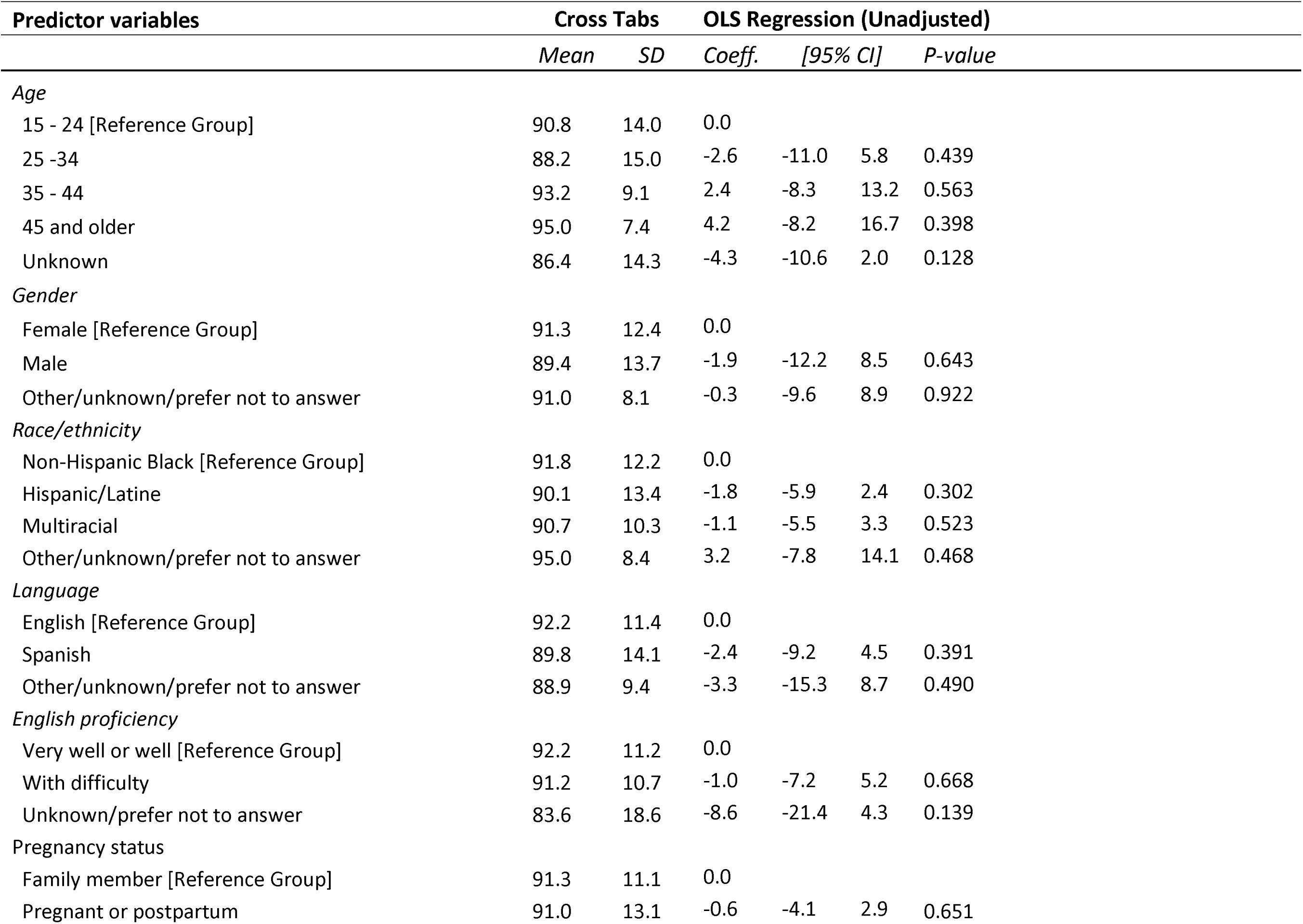
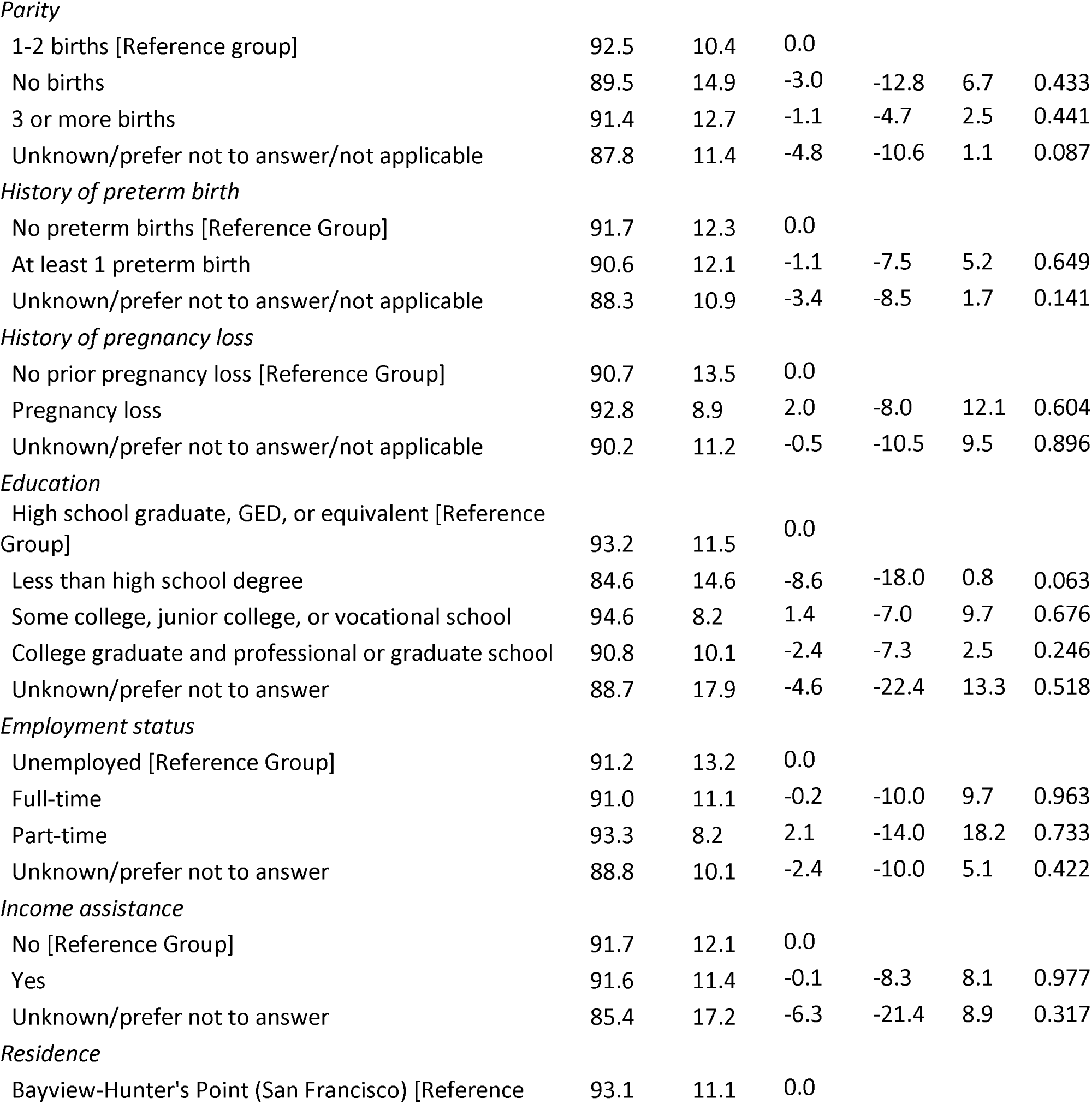
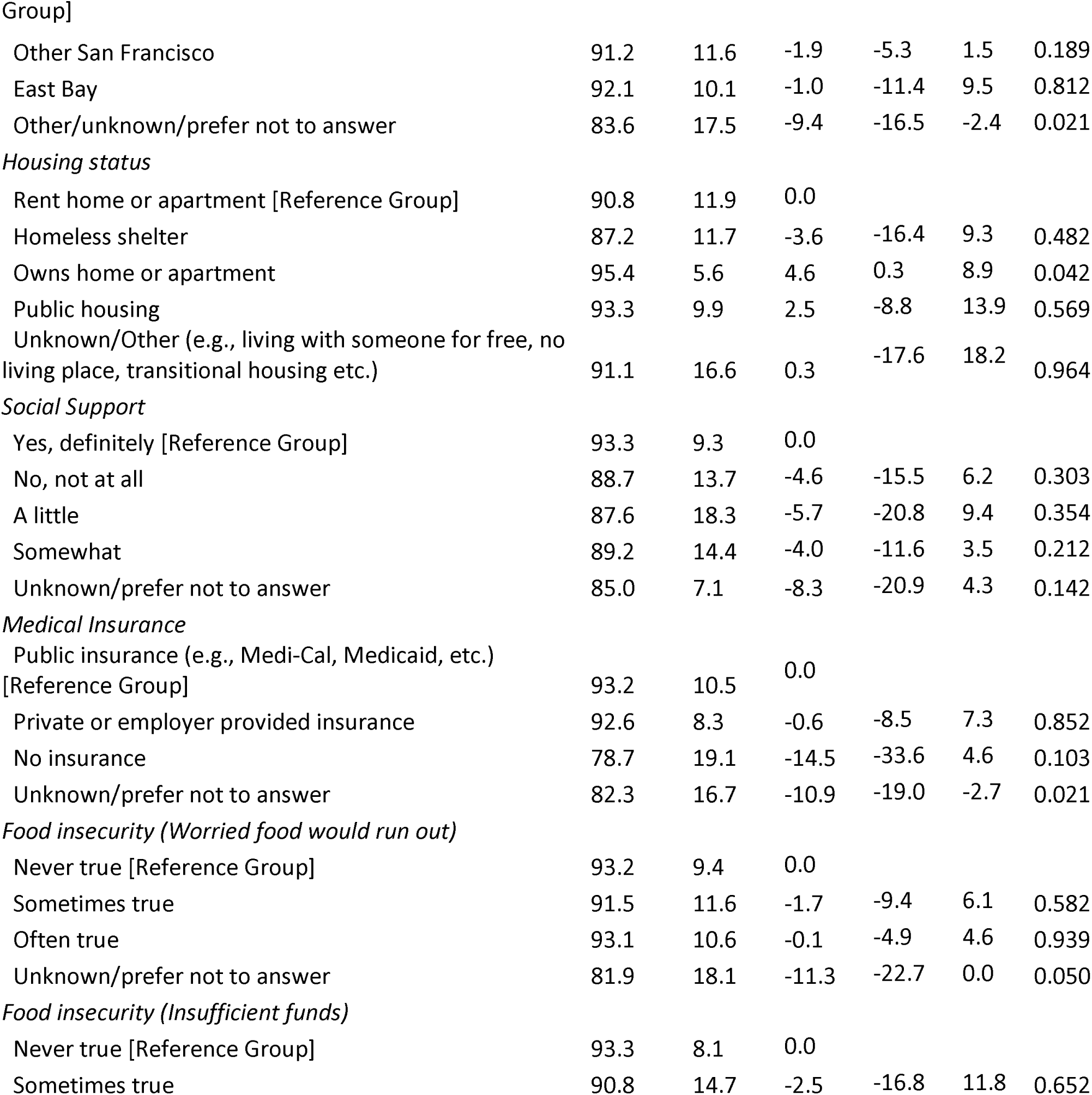
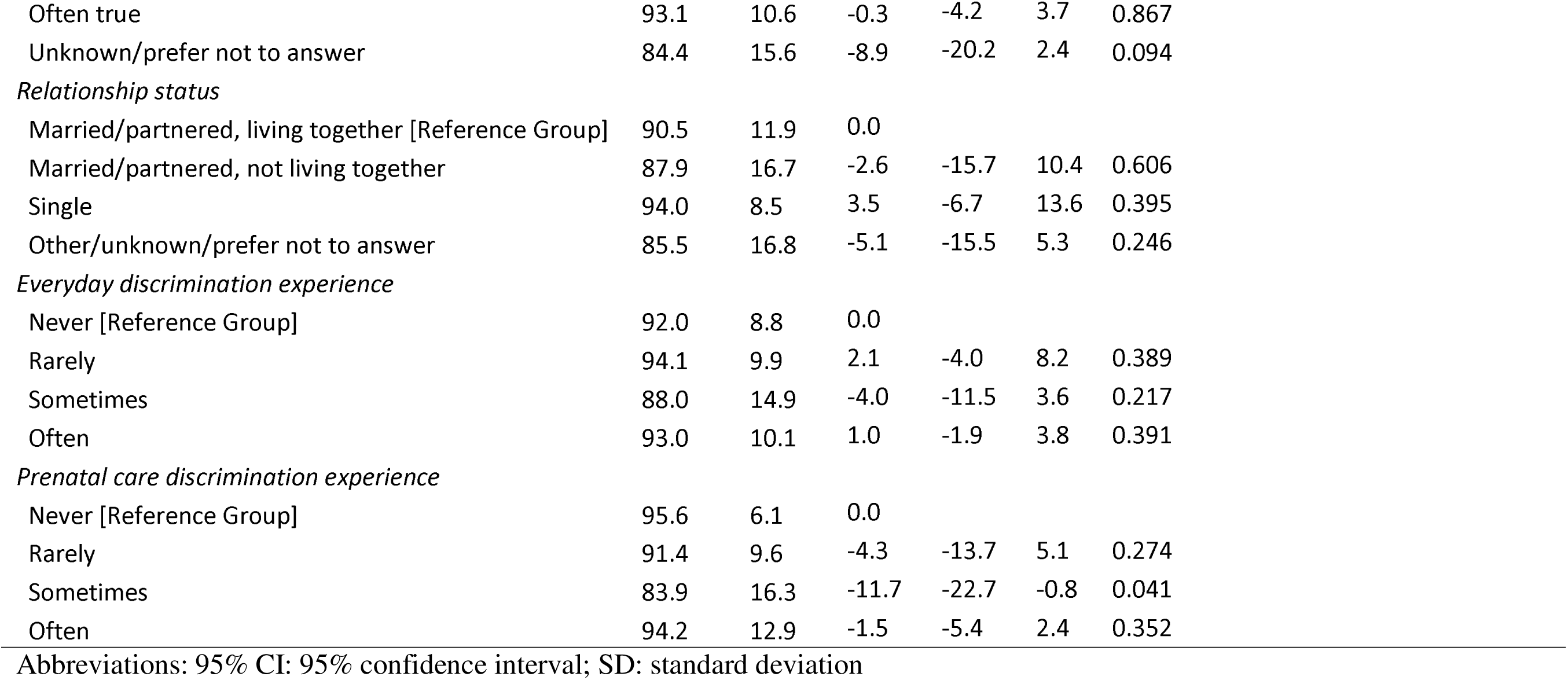
Bivariate statistics of sociodemographic characteristics, obstetric history, and care discrimination experiences for the full sample on the person-centered care score.

In the final multivariate model (Table 4), discrimination during prenatal encounters and medical insurance status were significantly associated with PCC. Participants who reported discrimination during prenatal care encounters on some occasions scored, on average, 10.9 points lower than those who reported no experiences of discrimination during prenatal care encounters (95% CI: -15.5 - -6.3). Participants without medical insurance scored, on average, 12.5 points lower than those with public insurance (95% CI: -22.1 - -2.9).

**Table 4.** Mixed-effects multivariate linear regression analysis of select predictors on the person-centered care score.

| Predictor variables | Coeff. | [95% CI] |  | p-value |
| --- | --- | --- | --- | --- |
| Residence |  |  |  |  |
| Bayview-Hunter's Point (San Francisco) [Reference Group] | 0.0 |  |  |  |
| Other San Francisco | -0.1 | -4.4 | 4.2 | 0.966 |
| East Bay | 4.5 | -3.8 | 12.9 | 0.288 |
| Other/unknown/prefer not to answer | -4.3 | -11.7 | 3.1 | 0.251 |
| Housing status |  |  |  |  |
| Rent home or apartment [Reference Group] | 0.0 |  |  |  |
| Homeless shelter | -3.4 | -9.7 | 3.0 | 0.297 |
| Owns home or apartment | 3.2 | -4.4 | 10.8 | 0.404 |
| Public housing | 2.5 | -3.0 | 8.0 | 0.377 |
| Unknown/Other (e.g., living with someone for free, no living place, transitional housing etc.) | 3.2 | -2.3 | 8.7 | 0.253 |
| Medical Insurance |  |  |  |  |
| Public insurance (e.g., Medi-Cal, Medicaid, etc.) [Reference Group] | 0.0 |  |  |  |
| Private or employer provided insurance | -0.5 | -6.8 | 5.8 | 0.870 |
| No insurance | -12.5 | -22.1 | -2.9 | 0.010 |
| Unknown/prefer not to answer | -8.7 | -15.1 | -2.3 | 0.007 |
| Prenatal care discrimination experience |  |  |  |  |
| Never [Reference Group] | 0.0 |  |  |  |
| Rarely | -3.9 | -9.0 | 1.3 | 0.139 |
| Sometimes | -10.9 | -15.5 | -6.3 | 0.000 |
| Often | -2.0 | -9.6 | 5.7 | 0.613 |
| Mean dependent var | 91.18 |  |  |  |
| Number of obs | 113 |  |  |  |
| Prob > chi2 | 0.000 |  |  |  |
| SD dependent var | 12.125 |  |  |  |
| Chi-square | 51.76 |  |  |  |
| Akaike crit. (AIC) | 872.99 |  |  |  |

Abbreviations: 95% CI: 95% confidence interval

#### Sensitivity analyses

The sensitivity analysis of the PCC score, in which missing and duplicate responses were excluded, resulted in an equivalent standardized average PCC score of 91.4 (SD = 11.4) for the main sample (*N* = 87), 92.0 (SD = 11.0) for pregnant and postpartum participants (N = 47) and 90.7 (SD = 12.0) for family members (N = 40).

### Qualitative Findings

#### Overview of qualitative findings

The qualitative data confirmed the high quantitative PCC scores. Overall, pregnant and postpartum people and their families found PV events to be person-centered, with a few notable exceptions. We expand on themes that emerged from the analysis organized by the PCC domains:

##### Responsive and supportive care

Participants stated that PV events were very supportive, attributing this to a welcoming environment and provider attentiveness. Participants also noted that providers were responsive to their needs by being attentive and going above and beyond, such as facilitating warm handoffs to other providers, making accessing services easier.

### Welcoming environment

Providers fostered a welcoming environment that enabled participants to seek out the care they needed, as they reported feeling warmth from service providers and were uplifted by it.

> “Just knowing that you are welcome somewhere is supportive – a feeling that you know, whether it’s for you or not. Just to hear that from someone as you pass their booth is very important. Everybody was pretty much in a good mood […] and people partook in what they were interested in.” —Multiracial family member, 45 years old and above

> “[People were] just welcoming. And, when they would see my little one, [they would ask] ‘Oh. Hey, how’s your [son]’ – you know, addressing him, asking how old he was, you know, when did I have him. Everyone was just warm and concerned, you know, warm and showed concern […] I felt like everyone was there and was just super supportive.”

> *—*Black postpartum participant, 45 years old and above

### Provider attentiveness

Participants reported that providers were highly attentive to their needs and sought ways to support them. One participant remarked on an interaction she had with one of the providers, who, after seeing her holding a flyer for childcare, spent the time to assess her needs for childcare and provide her resources related to childcare:

> “I felt like he was really paying attention to me even though that wasn’t necessarily the reason why I went over there. But he saw that I was reading, and he really wanted to make sure that I understood what kind of resources I had with childcare.” —Pregnant participant, less than 30 years old, race/ethnicity unknown

Another participant reflected on her appreciation of a provider taking the time to explain a procedure and address the participant’s concerns fully, ensuring the participant felt comfortable:

> “For example, the first time I went, I went into acupuncture, and it was a good service even though it was my first time in life. I had never had an acupuncture session. But I told the provider, that I was a little scared because I was pregnant, but the person told me nothing happens, it’s totally safe, it will help you and your baby. It’s like therapy, and the place was very comfortable, and she was always present; at the moment, I didn’t want to continue with acupuncture; she would stop and remove them immediately. She was very attentive!” —Latine postpartum participant, 30-44 years old

Others remarked that the staff and providers took the time to first understand their needs before discussing available resources. Once their needs were assessed, participants were personally guided to the booth or the person who would best be able to provide them with support and resources.

> “Yes, I [felt supported] because they listened first. They asked me what was my needs, what was going on in my life, you know, how was my health doing, before they gave me any information.” —Black pregnant participant, 30-44 years old

Participants who came on behalf of their pregnant or postpartum family members likened the support PV events provided them to prioritizeing the welfare of their family:

> “I felt supported. I felt like they [PV providers] genuinely cared about the welfare and the well-being of my family and me. And they wanted to make sure that I was well-informed and, with her [my pregnant daughter] being so young, that I can well inform her about this whole process. So, I have only excellent and great things to say about Pop-Up Village.”

> *—*Black family member, 45 years old and above

### Patient visibility

Participants credited provider responsiveness to feeling acknowledged, seen, and understood, as the providers were committed to delivering quality care. This was reflected in participant comments around model attentiveness to the needs of pregnant participants and provider responsiveness through keen observance of non-verbal cues suggesting that help was needed, and prompt response.

> “I felt very acknowledged as a pregnant woman. That was really positive. Just knowing that, oh, okay, they really set this up to serve this population, and here they are actively reaching out to someone who is pregnant and making sure that I am taken care of.”

> *—*Pregnant participant, less than 30 years old, race/ethnicity unknown

> “They were very understandable and met my needs. And, when they saw that I had like a lot of items in my hand, they went and offered and asked to grab bags for me so I could put them inside because I was by myself.” —Multiracial pregnant participant, 30-44 years old

### Warm handoffs

Participants expressed their appreciation for providers’ commitment to go above and beyond to ensure they received the necessary care and resources. This was evident in the providers’ proactive efforts to reach out to participants during events, answer any inquiries, and even coordinate with other service providers when needed. If a provider was unsure about an answer or how to locate the required resources or care for a participant, they sought out the information by connecting them with other PV providers who were more equipped to meet their needs.

> “They would walk up to me first and take initiative. I didn’t feel like I had to chase anybody down. As soon as I made eye contact with them, they would come over and ask, ‘Do you have a question?’ […] If they didn’t know the answer to something, they would find it out for me.” —Latine postpartum participant, 30-44 years old

> “When I had a question I asked, they answered if they could. If they didn’t, they’d try to find out. If they didn’t know it, they went somewhere to find out […] That’s what we needed. ‘If we don’t know, we can go somewhere or call somebody who probably knows more than we do and ask them.’ That’s what I enjoyed about it” —Black family member, 45 years old and above

> “If I needed assistance with anything [they would take] my information down and get me to the right person or to the right program. And they also directed me to, like, areas that I didn’t know where the booths were. So, they would actually take me to the booth and introduce me to the next person.” —Multiracial pregnant participant, 30-44 years old

#### Lack of provider follow-up

When participants were asked about negative experiences, they noted that lack of provider follow-up after the events negatively influenced their perceptions of the PCC they received. They mentioned that while having all necessary information consolidated in one location facilitates easier access to services, it can also feel overwhelming at times. As a result, participants desired closer follow-up with the services they accessed at PV after the events. However, some noted that they did not receive any follow-up from service providers after leaving the event.

> “I had a lot of folks come and talk to me about different things […] I expect to get a phone call or email because I feel like it was such a good resource […] I felt everywhere I went, everyone did talk and explain the resources, and I hope it’s like a follow-up phone call or email, just because it was so much information.” —Black pregnant participant, 30-44 years old

> “Well, with [X organization] my experience was good up until they didn’t call me back. They didn’t follow up. So, I was like they were not accountable for their word. So, I felt like that was pointless because that was the main focus of why I wanted to go. And that’s why my doctor encouraged me to go, to get that interaction with them and to set that all up with a doula.” —Multiracial pregnant participant, less than 30 years old

> “Once again, I only went to two or three places [organization booths], but one of them, I don’t get any calls yet. I don’t know if they open today or how it works with the system, but they’re supposed to be calling me; I don’t know when. Hopefully soon.” —Latine pregnant participant, 30-44 years old

#### Dignified and Respectful care

Participants noted that providers were respectful during care encounters, attributing their positive perceptions to providers’ intentionality to support and uplift them and their inviting and friendly demeanor.

### Provider intentionality

Participants reported feeling respected by the providers, who were intentional in their support. They particularly appreciated that the providers offered support to everyone, regardless of their birthing status, and approached their inquiries in a non-judgmental, respectful, and dignified manner to better understand and meet their needs.

> “I like that their intentions are good. When they approach you, their intentions are to help people, especially if they’re pregnant or not pregnant, or just giving birth.” —Latine postpartum participant, less than 30 years old

> “[The provider] asked a lot of clarifying questions. I thought that was really respectful. He didn’t assume I had childcare. He didn’t assume I could afford it or couldn’t afford it. He didn’t even assume I was looking for it. He asked questions in a way where I felt like he just really wanted to see how he could help me.” —Pregnant participant, less than 30 years old, race/ethnicity unknown

Participants also noted providers’ intention to uplift participants during care encounters. For example, a participant remarked on how empowering it felt to be respected, valued, and uplifted in her conversation with a provider. Such interactions permitted participants to be able to learn and glean more information from the resources available to them.

> “They never said anything that was derogatory or nothing that would put you down. Everything they said was lifting [you] up. You learned more. That made me feel good … The whole time I was there, I was up; never down. I was always lifted up and I liked that.” —Black family member, 45 years old and above

> “…These people [providers] were very engaged. They’ll hear you out. They’re very polite. If we accidentally start a sentence at the same time, they’ll yield and they’ll let you go first … It seems like they just have a heart for doing what they’re doing. It was very women-empowered, I believe. Just very gentle and soft.” —Latine postpartum participant, 30-44 years old

### Provider friendliness

Participants shared that they felt respected by the providers due to their friendly, welcoming, and genuine demeanor, which made participants feel at home and comfortable receiving services.

> “Well, I know that one of the girls there at the doula booth actually took some shifts here at X organization. And I recognized her. And she took the time to come eat lunch with me. We both went to get the food truck, and we caught up. But she gave me some vitamins. Well, because she knew I already had the baby.” —Latine postpartum participant, 30-44 years old

> “The [providers] were very helpful. I knew a couple of them. They like to interact with [my daughter]. They always ask about [her]. When you walk up into a spot and somebody’s like, “How old is she?” […] And people are genuinely interested in me, and that’s always nice. […] I grew up in Texas. So, in Texas, people are, like, you’re driving down a back country road, and people wave at you whether they know you or not, and [PV] kind of reminded me of that. Kind of like just very hometowny. Like, “How you doing, neighbor?” “Okay, yeah, I’m good.” “Good, all right!” Yeah, very supportive.”

> *—*Latine postpartum participant, 30-44 years old

#### Communication and autonomy

Participants noted that the providers communicated effectively with them, highlighting their knowledge, inclusivity, sincerity, and ability to ensure that information was clearly understood. However, a few participants shared that they received unclear information regarding the range of services available to them, with Spanish-speaking participants in particular expressing that the language barrier posed a challenge in communicating with providers.

### Provider knowledge

Providers’ awareness of available resources and their knowledge and understanding of their skillset or the resources their respective organizations held was important to participants. As a result, participants more easily trusted the advice and information these providers gave them.

> “Every last one of them was very detailed with what they were sharing with the general public. They were very well knowledgeable of what they wanted us to learn and have a takeaway so we can have a clear understanding what their programs represented.”

> *—*Black family member, 45 years old and above

> “[Providers at] each booth, [were] pretty much all in the medical profession. So, they’re really knowledgeable of what they do, what the service they provide, and if they are unsure, they had a colleague there to answer a question, or they can direct you into or to the right booth for information.” —M7503, Black pregnant participant, 30-44 years old

### Inclusivity

Communication with providers was viewed positively and empowering as participants felt included in conversations. Some participant responses included:

> “I’m pregnant myself, so I felt like people were really looking out for me and trying to start conversations, letting me know what is out there […] Whenever I stopped to look at what they offered, they included me in the conversation or tried to start a conversation with me to make sure I understood what they were offering. Yes. It was pretty positive.” —pregnant participant, less than 30 years old, race/ethnicity unknown

### Understanding of information

Participants felt that providers provided sufficient information and took the time to answer all of their questions. They highlighted the providers’ emphasis placed on sharing clear and in-depth information, even if that meant repeating responses multiple times for some participants. Some participant responses included:

> I think they answered, really, all my questions. Even though I didn’t ask a question, all the questions that I wanted to ask, they were answered when they were letting me know about the programs, like, for example, the Medi-Cal – I mean the food stamps and like the getting on like – stuff like that. I didn’t really have to ask a lot of questions. Because, when they were explaining it, my questions were getting answered. —Multiracial pregnant participant, less than 30 years old

> “When I didn’t understand a question, they would actually sit there and break it down to the point where I understand. Like, sometimes, I can comprehend [the information]. But, when I’m overwhelmed, I, you know, tend to ask to repeat it again. So, they had no problem repeating themselves.” —Multiracial pregnant participant, 30-44 years old

### Provider Genuity

Additionally, participants felt that providers truly cared about the participants they were communicating with, as they never felt rushed during their conversations and felt free to take as much time as necessary with each provider. For example, a family member appreciated the meaningful interactions she experienced with providers, highlighting how they took the time to engage in conversations until all her questions were thoroughly answered:

> “As long as I stood up there and was talking to them [the providers]. It might be 15 minutes. It might be 20 minutes. Until I felt like the questions I asked were answered. Then, I would go to somebody else and ask a question. They were really helpful. I enjoyed it very much.” —Black family member, 45 years old and above

> “Until the very end, until I left that place, they [the providers] were with me. They were just telling and explaining everything about the event. And then, they let me have my own time to see and to visit everything on my own. And then, they were waiting for me if I had any questions or any comments. Yeah. They really took care of me, really. Very good. Very, very good.” —Black pregnant participant, 30-44 years old

### Unclear messaging and language barriers

Some participants reported that they did not receive clear communication from providers regarding the full range of available services, which led to confusion and frustration about what services were offered and made it harder to access services.

> “So when I went [to PV], and I see just a bulletin and nobody, nobody like welcoming you, … I just had to ask around, and they told me, like, “Okay, you can get some information here in that first booth.” But I didn’t really get that good information; I was like, “How can I do [this]? What should I do here? […] There wasn’t like one, you know, one booth that they can kind of give details about everything, you have to go like one by one…” —Latine pregnant participant, 30-44 years old

> “I had a lot of information from some [providers] that is very useful to me, but I was also looking for services, specifically massage. So, when I went the first time, I was looking for massages for pregnant women. I know they exist, but economically I do not have [the money] to pay for a private one. So, I went there because I thought massages were offered to pregnant women. [… When] I went for the first time, I looked for it and did not find it. Last time I went […]and I did not find it. I contacted a provider, and when they were at Pop-up Village, they told me that they had the services, […] However, when I arrived, they told me that, right now, they were not doing it [providing those services] […] If you’re going to talk about a resource that you have, make sure it’s true.” —Latine postpartum participant, 30-44 years old

Other communication challenges reported were due to a lack of Spanish-speaking providers; some Spanish-speaking participants felt lost and struggled to convey their needs.

> “It was hard for me a little when I arrived to look for someone who spoke Spanish and then it is the only thing I would say if other people arrived as well as me. We know English, but we would like to speak more Spanish, as at the entrance, there should be someone who speaks Spanish.” —Latine family member, less than 30 years old

## DISCUSSION

In this mixed-methods study, we examined participant perspectives on the person-centeredness of the PV events, including its ability to prioritize participants’ values and needs while bridging gaps in health inequities. Our findings show that the PV events based on the Pregnancy Village model of meeting comprehensive needs through cross-sector care delivery were perceived to be person-centered by pregnant and postpartum people and their families. Through our qualitative findings, we delineate how the dimensions of PCC, including respectful care, communication and autonomy, and responsive and supportive care, were integral in shaping community members’ perceptions of PCC at PV events. Additionally, the findings highlighted the need for ongoing efforts to address consistent follow-up, clearer information delivery, and language concordance to optimize PCC within this model.

Both quantitative and qualitative data indicate that PV events were responsive to the needs and values of pregnant and postpartum individuals and their families while respectfully providing care. Notably, our sample reported higher PCC than other literature on healthcare experiences of Black and other minoritized individuals during pregnancy (27–30). This is likely because the model was intentionally designed to improve PCC, and there was significant work done with all participating providers around the core goals of the program: being responsive to the needs of participants, focusing on interpersonal communication, and fostering a supportive care environment where individuals feel heard and respected. Also, the iterative nature of the model, where feedback is continually gathered from individuals to inform improvement, aims to ensure that gaps in PCC are promptly addressed.

Highly valued provider behaviors that contributed to the person-centeredness of events included provider attentiveness, warm handoffs, and inclusivity. Participants valued provider recognition and response to participants’ non-verbal cues that indicated a need (attentiveness). How well providers respond to non-verbal cues is indicative of their empathy, which encompasses both cognitive and emotional elements, allowing individuals to recognize and respond to others’ spoken and unspoken needs and concerns, a critical aspect of PCC (31). Attentiveness may be particularly important for Black and other minoritized individuals who are less likely to seek help than their white counterparts due to fear of racial discrimination, legal status, and cultural stereotypes (e.g., the “Strong Black Woman” or “Superwoman” schema of suppressing one’s vulnerabilities and avoiding dependence) (32–34).

Warm handoffs, facilitated by service co-location, were similarly valued to connect individuals and their families with relevant resources, fostering a sense of empowerment and, thus, higher perceptions of PCC (35,36). This contrasts with Black and other minoritized people’s experiences accessing care in the health system and social service bureaucracy, which frequently operate in isolation, with limited communication and collaboration, creating obstacles to coordinated care (37–40). Finally, inclusivity— involving participants in discussions— was also highly valued. Successful provider-patient communication and engagement through active listening and open-ended questions to assess patient needs and preferences reduces the individual’s burden to advocate for themselves (41). Prior research indicates that minoritized individuals often struggle to advocate for themselves, with Black women being the least likely to do so in healthcare settings (42). Thus, actively involving individuals in dialogue and uplifting their viewpoints and lived experiences encourages open communication and promotes shared decision-making, empowering individuals in their care experience (43).

Despite the generally positive perceptions of PV events on PCC across domains, the few instances of poor PCC that had a negative impact on individuals’ experiences included a lack of provider follow-up, miscommunication, and language barriers. Lack of follow-up can lead to feeling abandoned and frustrated, with a diminished sense of control over health and well-being and the perception that the individual is not valued in their care (44). Lack of provider follow-up can also erode people’s trust in both providers and institutions, thus contributing to poorer health outcomes (45). This finding indicates that models designed to deliver collocated services need to prioritize effective follow-up processes to prevent individuals from feeling disempowered and unsupported around their health and well-being after events. Miscommunication and language barriers, partly due to fewer Spanish-speaking providers relative to Spanish-speaking participants, can cause frustration and compromise fundamental PCC principles of meeting individuals’ needs and preferences while safeguarding their autonomy. It can also result in individuals feeling ignored, misunderstood, and possibly not receiving appropriate care (46,47). For programs where priority populations include non-English speakers, an adequate number of language-concordant providers and individuals who have a deep cultural understanding and can serve as effective advocates are critical. Further, it is important to establish clear expectations and deliver information in a straightforward manner that is practical, relevant, and meaningful to individuals.

Finally, our finding that individuals who faced discrimination during their prenatal care interactions reported lower PCC compared to those who did not is worth noting. This is likely due to the intricate dynamics between discrimination and health, particularly discrimination-related vigilance, a coping strategy where individuals try to protect themselves from expected discrimination by being vigilant of their behavior and environment (48). In other words, just the expectation of experiencing discrimination can elevate stress levels during care encounters, which in turn can negatively impact the overall care experience (49). This finding suggests the need for providers to reflect the communities they serve and possess a cultural competence rooted in their professional training and shared lived experiences (50). Where providers do not reflect their patients, training on unconscious bias in provider behavior and potential patient responses due to prior experience and discrimination-related vigilance, rooted in cultural humility, is essential.

To our knowledge, the Pregnancy Village model of care is the first that focuses on improving perinatal outcomes by providing a space for accessing cross-sector services, uplifting and celebrating Black and other minoritized pregnant people, and fostering trust and relationship building between community and institutions (14), and our study provides an example of how such models may be evaluated. Notable strengths of the study include the mixed methods design, which allowed us to obtain quantitative scores from a larger sample and in-depth qualitative data to expand on the quantitative data. Also, the PCC scale used was adapted from a validated scale developed for the PV priority population (Black pregnant and postpartum people) (51).

Nonetheless, our findings should be considered in light of several limitations. We used convenience and purposeful sampling to recruit participants, which limits generalizability. The small sample size for the semi-structured interviews further limits generalizability. However, the purpose was to enhance understanding of the quantitative PCC scores and the consistency in the results, as well as achieve thematic saturation and support trustworthiness, which is the goal of qualitative methodologies. Finally, given that all data are self-reported, social desirability and recall bias are also potential limitations.

## CONCLUSION

We demonstrate that the Pregnancy Village model of bringing cross-sector organizations together to provide services as a community-based one-stop-shop is not only possible (14) but can be done in a person-centered manner that prioritizes individual needs, values, and preferences. Such a model may serve as an avenue to mitigate lack of access and adverse care experiences that continue to contribute to the disparities in maternal and infant morbidity and mortality in the United States. However, ensuring consistent provider follow-up with individuals, clear communication, and language concordance are key components for this event-based care delivery model to have optimal impact.

## ACKNOWLEDGEMENTS

We would like to thank all the organizations, staff, and volunteers who participated in the PV events, providing care and services to San Francisco’s underserved residents. We are also grateful to the research assistants for their valuable contributions to qualitative data analysis. Finally, we thank all PV participants, especially those who generously shared their experiences through the interviews.

## AUTHOR CONTRIBUTIONS

PCD: Investigation, validation, methodology, formal analysis, writing – original draft, writing – review and editing; OJO: Investigation, validation, methodology, project administration, data curation, formal analysis, writing - original draft, writing – review and editing; AJB and AME: Conceptualization, investigation, validation, methodology, data curation, writing – review and editing; MAN: Conceptualization, investigation, validation, methodology, resources, funding acquisition, supervision, writing – review and editing; KC: project administration, writing – review and editing; PAA: Conceptualization, investigation, validation, methodology, project administration, data curation, resources, funding acquisition, writing - original draft, writing – review and editing

## STATEMENTS AND DECLARATIONS

### Ethical considerations

We obtained ethics approval from the Institutional Review Board of the University of California, San Francisco (#20–32393).

### Consent to participate

All participants provided verbal informed consent prior to participating.

### Consent for publication

Not applicable.

### Data availability

The datasets generated during and/or analyzed during the current study are not publicly available due to privacy and ethical restrictions but are available from the corresponding author upon reasonable request.

## Footnotes

1 Not all individuals who are pregnant or have given birth identify as women. Where we use the term “individuals” for brevity, it should be understood to include “women and all other individuals who are pregnant or have given birth” (2).

2 Participants who attended multiple monthly PV events were allowed to complete one survey for each PV event that they attended. Fifteen participants completed the survey more than once. One participant completed the survey eight times, one completed the survey six times, two participants completed the survey three times, and eleven completed the survey twice.

